# Effect of underlying comorbidities on the infection and severity of COVID-19 in South Korea

**DOI:** 10.1101/2020.05.08.20095174

**Authors:** Wonjun Ji, Kyungmin Huh, Minsun Kang, Jinwook Hong, Gi Hwan Bae, Rugyeom Lee, Yewon Na, Hyoseon Choi, Seon Yeong Gong, Yoon-Hyeong Choi, Kwang-Pil Ko, Jeong-Soo Im, Jaehun Jung

**Author notes:** Wonjun Ji, Kyungmin Huh, and Minsun Kang^¶^ contributed equally to this work. **Correspondence to Jaehun Jung, MD, PhD**, Department of Preventive Medicine, Gachon University College of Medicine, 38-13, Dokjeom-ro 3, Incheon, Korea.

## Abstract

**Background:** The coronavirus disease (COVID-19) pandemic is an emerging threat worldwide. It is still unclear how comorbidities affect the risk of infection and severity of COVID-19.

**Methods:** A nationwide retrospective case-control study of 65,149 individuals, aged 18 years or older, whose medical cost for COVID-19 testing were claimed until April 8, 2020. The diagnosis of COVID-19 and severity of COVID-19 infection were identified from the reimbursement data using diagnosis codes and based on whether respiratory support was used, respectively. Odds ratios were estimated using multiple logistic regression, after adjusting for age, sex, region, healthcare utilization, and insurance status.

**Results:** The COVID-19 group (5,172 of 65,149) was younger and showed higher proportion of females. 5.6% (293 of 5,172) of COVID-19 cases were severe. The severe COVID-19 group had older patients and a higher male ratio than the non-severe group. Cushing syndrome (Odds ratio range (ORR) 2.059-2.358), chronic renal disease (ORR 1.292-1.604), anemia (OR 1.132), bone marrow dysfunction (ORR 1.471-1.645), and schizophrenia (ORR 1.287-1.556) showed significant association with infection of COVID-19. In terms of severity, diabetes (OR 1.417, 95% CI 1.047-1.917), hypertension (OR 1.378, 95% CI 1.008-1.883), heart failure (ORR 1.562-1.730), chronic lower respiratory disease (ORR 1.361-1.413), non-infectious lower digestive system disease (ORR 1.361-1.418), rheumatoid arthritis (ORR 1.865-1.908), substance use (ORR 2.790-2.848), and schizophrenia (ORR 3.434-3.833) were related with severe COVID-19.

**Conclusions:** We identified several comorbidities associated with COVID-19. Health care workers should be more careful when diagnosing and treating COVID-19 when the patient has the above-mentioned comorbidities.

**Take Home message:** Comorbidities that might be associated with COVID-19 infection and severe clinical course were identified, which could assist in formulating public health measures to mitigate the risk in groups with increased risk.

## INTRODUCTION

The novel coronavirus disease (COVID-19) caused by SARS-CoV-2, has rapidly emerged since December 2019. As of April 18, 2020, a total of 2,160,207 confirmed cases, including 146,088 deaths, have been reported worldwide[1]. Despite the introduction of social distancing to slow the spread in many countries, outbreak of COVID-19 has overwhelmed healthcare systems in several countries. Inadequate medical resources have warranted the identification of risk factors for the diagnosis and severity of COVID-19. Although early epidemiological studies from china reported the prevalence of comorbidities,[2, 3] and its association with disease severity[4, 5], the host factors which increase the risk of COVID-19 infection or its severity still need to be identified.

The national health insurance service of South Korea registered nearly all COVID-19 patients and test negative control within the national insurance system. Recently, the Health Insurance Review & Assessment Service (HIRA) of Korea agreed to the share invaluable national health insurance claims data related to COVID-19 for public health purposes. This data can be useful for the identification of underlying comorbidities which are associated with the diagnosis and severity of COVID-19.

We conducted a retrospective case-control study, examining the effect of various underlying comorbidities on the risk of infection, and severity of COVID-19 using data from the nationwide medical insurance claim database in South Korea.

## MATERIALS AND METHODS

### Data source

We extracted information from the insurance claims database of the HIRA of South Korea. The claims (Figure 1) are made with a special “public crisis” code (MT043) for every suspected case under the national insurance coverage. The reimbursement for confirmed cases are claimed with the Korean Standard Classification of Diseases and Causes of Death, 7th edition (KCD-7) codes, which is a modified version of the International Classification of Diseases and Related Health Problems, 10th edition (ICD-10), designated for COVID-19 (B34.2, B97.2, U18, U18.1, and U07.1). Thus, we identified all tested individuals within the national health insurance coverage using the code MT043. All subjects with KCD-7 codes for COVID-19 were categorized as reverse transcription polymerase chain reaction (RT-PCR) test positive cases when the diagnosis was confirmed using RT-PCR with respiratory specimens; the remaining subjects were categorized as controls (“RT-PCR test negative” controls). Moreover, severe cases were defined as patients with a diagnosis confirmed by an RT-PCR test, who had the claim data of oxygen therapy, mechanical ventilator, extracorporeal membrane oxygenation, and cardiopulmonary resuscitation. The remaining laboratory confirmed subjects were categorized as non-severe cases (Figure 2).

**Figure 1.**
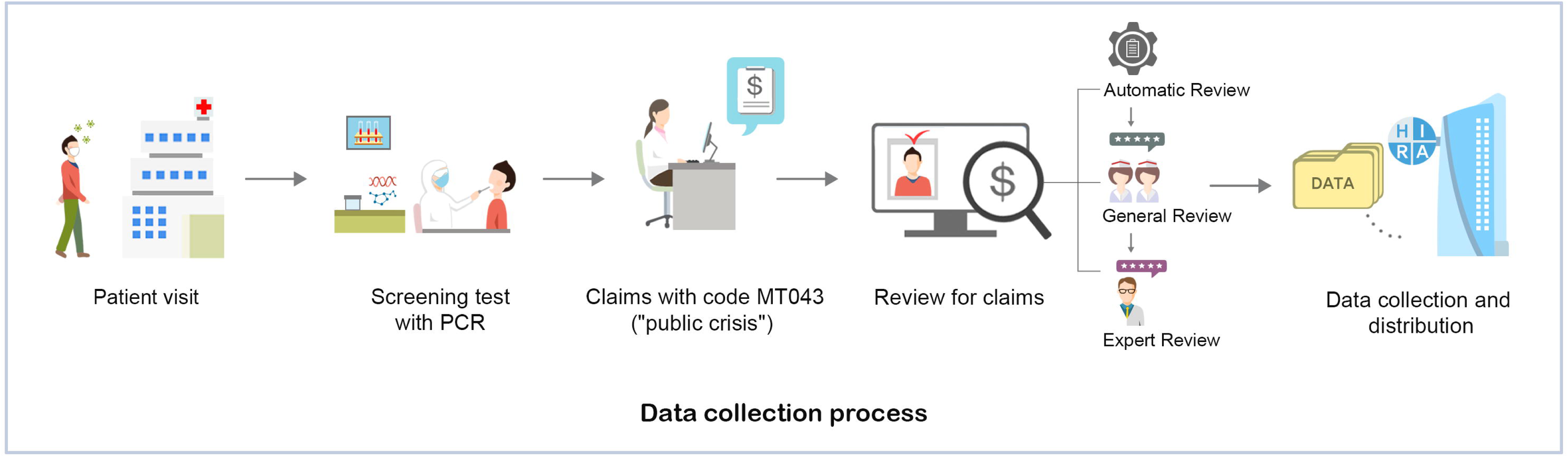
Overview of national health insurance claims data from Health Insurance Review & Assessment Service of Korea.

**Figure 2.**
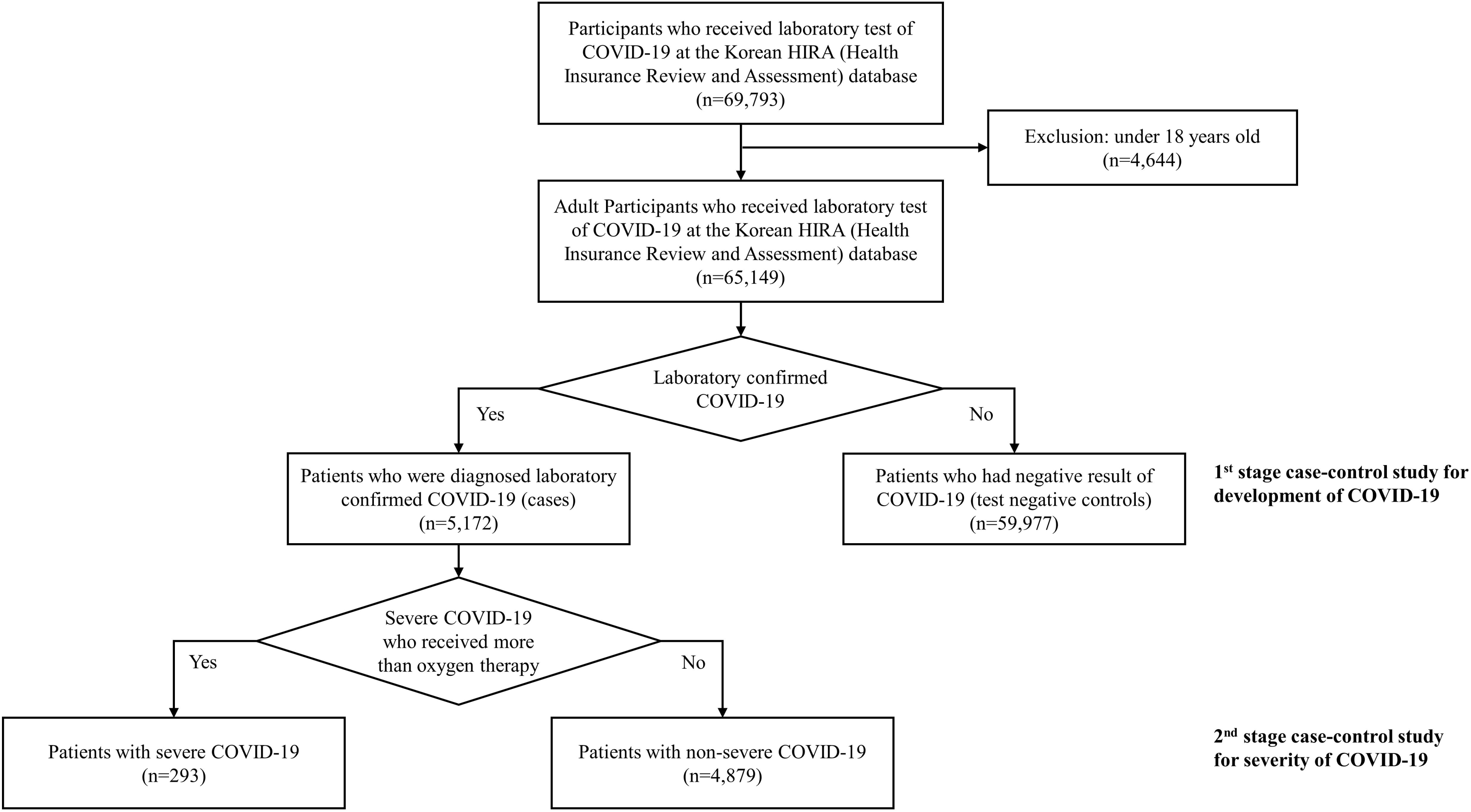
Flow chart of selecting process for study participants.

### Study design and definitions

This was a two staged retrospective case control study to evaluate the underlying comorbidities associated with diagnosis and severity of COVID-19. First, we examined the underlying comorbidities associated with the diagnosis of COVID-19 between laboratory confirmed cases and test negative controls. Also, the underlying comorbidities associated with severity of COVID-19 were evaluated and compared between the severe and non-severe confirmed cases. Underlying comorbidities were defined as the reimbursement for ≥2 times of KCD-7 code of study diseases, within 5 years prior to the test for COVID-19. The disease of interest was selected from the list of ICD-10 mapping tree, with reports of possible association with SARS-CoV-2 in previous epidemiologic studies, and those with theoretical concerns for increased risk (Table S1). Two authors (W. J. and K. H.) reviewed literature and selected the diseases of interest, and disagreement was arbitrated by the third author (J. J.). There were 56 categories in the disease group (Table S2). The location of the medical institution where the patients had been treated was identified to control a substantially higher risk of community acquired infection. Daegu city and Gyeongsangbuk-do province (DG), had a large regional outbreak and many cases did not have any identifiable trace[6]. A subgroup analysis was conducted for DG area and outside of DG area to identify the risk factors related to community outbreaks. Also, Charlson comorbidity index (CCI) was calculated as previously described (Table S3)[7]. Healthcare utilization was evaluated by the number of hospitalizations, the number of outpatient visits, and the number of emergency room visits within one year prior to tests for COVID-19. A subgroup analysis was also performed excluding patients with hospitalization over 30 days. This was done to avoid the confounding effect of long-term health care utilization.

### Statistical analysis

The baseline characteristics of cases and controls were compared using χ^2^ test and Student’s *t* test, as appropriate. The prevalence of comorbidities was compared using logistic regression models which adjusted with sex, age, residence, CCI, and healthcare utilization as covariates. To mitigate the risk of quasi-separation and overfitting, we performed two different type of multivariable analysis. The multivariable model 1, included one comorbidity at a time with all other covariates, constructing individual models for each comorbidity; the multivariable model 2, included all comorbidities and other covariates in a single model. We used the significance threshold of p<0.05, and all tests were two-tailed. SAS version 9.4 (SAS Institute Inc., Cary, NC, USA) was used for the analyses. This study was approved by the Institutional Review Board of the Gil Medical Center, Gachon University College of Medicine which provided a waiver of consent (GFIRB2020-134).

## RESULTS

### Baseline demographics between case and test negative controls

At the data cut-off time, on April 8, 2020, a total of 65,149 individuals aged ≥18 years who underwent laboratory tests for COVID-19 in South Korea were identified and analyzed. The proportion of females was slightly higher (2,883 of 5,172; 55.7%) than that of males in the case group. Few COVID-19 patients (1,335 of 5,172; 25.5%) were treated in medical facilities in the DG area, while 74.2% of confirmed cases, treated outside of the DG area. The control group was older and showed higher healthcare utilization within 1 year before undergoing the laboratory test for COVID-19. Overall, the case group showed lower prevalence than the control group except for glomerular disease (Table 1).

**Table 1.** Baseline demographics and Prevalence of comorbidities between case and test negative controls.

| Variables | Cases<br>(N=5,172) | Controls<br>(N=59,977) | P-value |
| --- | --- | --- | --- |
| **Demographic characteristics** |  |  |  |
| Sex, Male, n (%) | 2,289 (44.3) | 29,224 (48.7) | <.0001 |
| Age, median (range) | 42 (18-100) | 45 (18-110) | <.0001 |
| 18-50, n (%) | 3,214 (62.1) | 33,544 (55.9) | <.0001 |
| 50-65, n (%) | 1,144 (22.1) | 11,886 (19.8) |  |
| 65-80, n (%) | 608 (11.8) | 9,000 (15.0) |  |
| ≥80, n (%) | 206 (4.0) | 5,547 (9.2) |  |
| **Residence** |  |  |  |
| DG area, n (%) | 1,335 (25.8) | 7,553 (12.6) | <.0001 |
| Except DG, n (%) | 3,837 (74.2) | 52,424 (87.4) |  |
| Charlson comorbidity index, mean (SD) | 1.34 (1.9) | 1.75 (2.3) | <.0001 |
| **Healthcare utilization within 1 years before diagnosis of COVID-19** |  |  |  |
| Number of hospitalization, mean (SD) | 0.49 (1.6) | 0.93 (2.4) | <.0001 |
| Number of hospitalization over 30 days, mean (SD) | 0.01 (0.1) | 0.03 (0.2) | <.0001 |
| Number of outpatient visit, mean (SD) | 23.67 (34.1) | 24.83 (31.6) | 0.0175 |
| Number of ED visit, mean (SD) | 0.27 (0.9) | 0.52 (1.9) | <.0001 |
| Medical Aids, n (%) | 287 (5.5) | 3,434 (5.7) | 0.5999 |
| **Underlying Diseases** |  |  |  |
| **Endocrinopathy** |  |  |  |
| Diabetes | 713 (13.8) | 10,886 (18.2) | <.0001 |
| Thyroid disease | 325 (6.3) | 3,862 (6.4) | 0.6621 |
| Cushing syndrome | 10 (0.2) | 75 (0.1) | 0.1917 |
| Osteoporosis | 406 (7.8) | 6,014 (10.0) | <.0001 |
| **Cardiac disease** |  |  |  |
| Isolated Hypertension | 1,126 (21.8) | 17,613 (29.4) | <.0001 |
| Ischemic heart disease | 297 (5.7) | 5,303 (8.8) | <.0001 |
| Heart failure and cardiomyopathy | 217 (4.2) | 3,868 (6.4) | <.0001 |
| Valvular heart disease | 30 (0.6) | 565 (0.9) | 0.0086 |
| Cardiac arrhythmia | 194 (3.8) | 3,278 (5.5) | <.0001 |
| **Chronic respiratory disease** |  |  |  |
| Chronic upper respiratory disease | 3,628 (70.1) | 42,446 (70.8) | 0.3444 |
| Chronic lower respiratory disease | 1,532 (29.6) | 21,624 (36.1) | <.0001 |
| Environmental lung disease | 10 (0.2) | 327 (0.5) | 0.0007 |
| Interstitial lung disease | 18 (0.3) | 429 (0.7) | 0.0021 |
| Chronic respiratory failure and | 4 (0.1) | 72 (0.1) | 0.3880 |
| Diaphragm palsy |  |  |  |
| Pulmonary vascular disease | 15 (0.3) | 336 (0.6) | 0.0109 |
| **Renal disease and ESRD** |  |  |  |
| Hypertensive renal disease | 61 (1.2) | 527 (0.9) | 0.0282 |
| Glomerular disease | 121 (2.3) | 1,169 (1.9) | 0.0531 |
| Renal tubule-interstitial disease | 31 (0.6) | 535 (0.9) | 0.0296 |
| History of Acute Renal failure | 27 (0.5) | 588 (1.0) | 0.0011 |
| Chronic renal failure and ESRD | 234 (4.5) | 2,478 (4.1) | 0.1748 |
| Urolithiasis | 86 (1.7) | 1,085 (1.8) | 0.4476 |
| **Viral hepatitis and Chronic liver disease** |  |  |  |
| HBV, acute and chronic | 100 (1.9) | 1,226 (2.0) | 0.5888 |
| HCV, acute and chronic | 13 (0.3) | 278 (0.5) | 0.0281 |
| Non-B, non-C hepatitis | 450 (8.7) | 6,337 (10.6) | <.0001 |
| Liver cirrhosis | 40 (0.8) | 1,055 (1.8) | <.0001 |
| Hepatic failure | 7 (0.1) | 188 (0.3) | 0.0245 |
| **Disease of digestive system** |  |  |  |
| Non-infectious disease of Upper Digestive system | 4,804 (92.9) | 56,086 (93.5) | 0.0797 |
| Non-infectious disease of Lower Digestive system | 1,838 (35.5) | 24,740 (41.2) | <.0001 |
| Pancreatic disease | 57 (1.1) | 1,354 (2.3) | <.0001 |
| Biliary disease | 124 (2.4) | 2,488 (4.1) | <.0001 |
| **Chronic neurologic disease** |  |  |  |
| Systemic atrophy | 5 (0.1) | 98 (0.2) | 0.2465 |
| Parkinsonism and movement disorder | 209 (4.0) | 2,637 (4.4) | 0.2298 |
| Alzheimer and degenerative disease | 134 (2.6) | 2,559 (4.3) | <.0001 |
| Multiple sclerosis | 1 (0.0) | 66 (0.1) | 0.0508 |
| Epilepsy | 97 (1.9) | 1,955 (3.3) | <.0001 |
| Transient cerebral ischemia | 349 (6.7) | 6,306 (10.5) | <.0001 |
| Stroke, Cerebral hemorrhage |  |  |  |
| Dementia | 172 (3.3) | 3,771 (6.3) | <.0001 |
| **Malignancy** |  |  |  |
| Solid organ, except respiratory, thyroid | 247 (4.8) | 4,779 (8.0) | <.0001 |
| Respiratory tract | 33 (0.6) | 937 (1.6) | <.0001 |
| Thyroid cancer | 67 (1.3) | 646 (1.1) | 0.1476 |
| Hematologic | 17 (0.3) | 489 (0.8) | 0.0001 |
| **Rheumatologic disease** |  |  |  |
| Rheumatoid arthritis | 117 (2.3) | 1,923 (3.2) | 0.0002 |
| SLE | 0 (0.0) | 0 (0.0) |  |
| Systemic connective tissue disease | 51 (1.0) | 694 (1.2) | 0.2670 |
| **Hematologic disease** |  |  |  |
| Anemia | 590 (11.4) | 7,531 (12.6) | 0.0164 |
| Coagulopathy | 42 (0.8) | 921 (1.5) | <.0001 |
| Bone marrow dysfunction | 45 (0.9) | 630 (1.1) | 0.2191 |
| **Obesity** | 2 (0.0) | 122 (0.2) | 0.0091 |
| **Nutritional deficiency** | 239 (4.6) | 4,077 (6.8) | <.0001 |
| **Mental and Behavioral disorders** |  |  |  |
| Mental disorder of Substance use | 36 (0.7) | 685 (1.1) | 0.0033 |
| Schizophrenia | 118 (2.3) | 1,270 (2.1) | 0.4331 |
| Mood disorder | 588 (11.4) | 8,956 (14.9) | <.0001 |
| Neurosis | 820 (15.9) | 11,112 (18.5) | <.0001 |
| Personality disorder | 11 (0.2) | 126 (0.2) | 0.9687 |
| Mental retardation, Development disorder | 17 (0.3) | 143 (0.2) | 0.2082 |
| **Immune deficiency, HIV infection** | 9 (0.2) | 99 (0.2) | 0.8793 |

IQR, Interquartile range. DG, Daegu city and Gyeongsangbuk-do province area. ESRD, end-stage renal disease. HBV, hepatitis B virus. HCV, hepatitis C virus. SLE, systemic lupus erythematosus. HIV, human immunodeficiency virus.

### Comorbidities associated with diagnosis of COVID-19

#### Result of Overall analysis

Figure 3 and Table S4 represented the Odds ratios (ORs) for the diagnosis of COVID-19 according to the 56 categories of comorbidities. Hypertensive renal disease was associated with an increased risk of COVID-19 in a univariate analysis. In terms of multivariable analysis, Cushing syndrome (OR range; ORR 2.059-2.358), glomerular disease (ORR 1.366-1.445), chronic renal failure (CRF) and end stage renal disease (ESRD) (ORR 1.292-1.493), bone marrow dysfunction (ORR 1.471-1.645), and schizophrenia (ORR 1.287-1.556) showed an increased risk in the whole study population (Table S4) and in the subgroup that excluded long term health care utilizers (Table S5). Anemia (OR 1.132, 95% confidential interval; CI 1.021-1.254) showed an increased risk only in multivariable analysis with single disease category, while hypertensive renal disease (OR 1.604, 95% CI 1.206-2.133) showed an increased risk not only in multivariable analysis with single disease category, but also in subgroup analysis of multivariable analysis with whole disease category (Table S5). Isolated hypertension (ORR 0.840-0.867), chronic lower respiratory disease (ORR 0.856-0.876), liver cirrhosis (ORR 0.633-0.663), lower digestive system diseases (ORR 0.892-0.926), pancreatic disease (ORR 0.718-0.746), biliary disease (ORR 0.782-0.825), epilepsy (ORR 0.747-0.765), dementia (ORR 0.764-0.809), obesity (ORR 0.192-0.195), and mood disorder (ORR 0.859-0.896) showed a decreased risk of COVID-19 in multivariable analysis.

**Figure 3.**
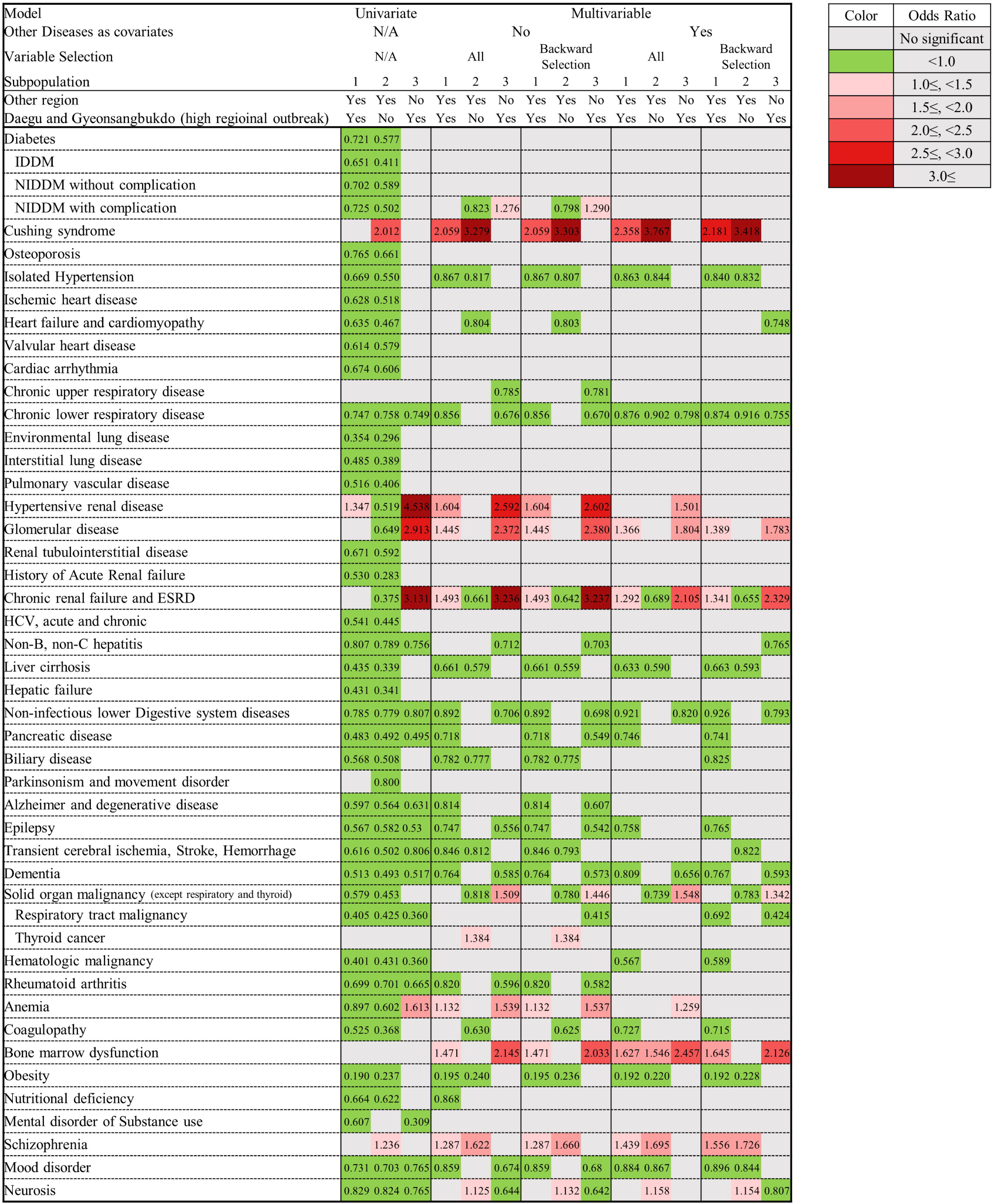
Analysis of relationship between comorbidities and infection of COVID-19.

##### Analysis of subpopulation which excluded high regional outbreak area (DG area)

Cushing syndrome (ORR 3.279-3.767) and schizophrenia (ORR 1.622-1.726) were associated with an increased risk of COVID-19 in univariate and multivariable analysis. And neurosis (ORR 1.125-1.158) showed an increased risk of COVID-19 in multivariable analysis (Figure 3, Table S6). Although thyroid cancer (OR 1.384, 95% CI 1.020-1.878) was shown an increased risk in multivariable model 1 with single disease category, there was no significance in multivariable model 2 with whole disease category. Isolated hypertension (ORR 0.807-0.844), chronic lower respiratory diseases (ORR 0.902-0.916), CRF and ESRD (ORR 0.661-0.689), liver cirrhosis (ORR 0.559-0.593), cerebrovascular diseases (ORR 0.793-0.822), solid organ malignancy (ORR 0.739-0.818), and obesity (ORR 0.220-0.240) showed a decreased risk of COVID-19 in multivariable analysis. This pattern was consistent in the subgroup which excluded long term health care utilizers (Table S7).

#### Analysis of subpopulation in high regional outbreak area (DG area)

This subpopulation analysis was conducted to identify the risk factors for high regional outbreak. Hypertensive renal disease (ORR 1.501-2.602), glomerular disease (ORR 1.783-2.380), CRF and ESRD (ORR 2.105-3.237), and anemia (ORR 1.259-1.539) were associated with an increased risk of COVID-19 outbreak in univariate and multivariable analysis. Solid organ malignancy (ORR 1.342-1.548) and bone marrow dysfunction (ORR 2.033-2.457) was associated with an increased risk of COVID-19 only in multivariable analysis (Figure 3, Table S8). The patients who had non-insulin dependent diabetes with complication (OR 1.267-1.290) was associated with an increased risk of presence of COVID-19 in multivariable model 1. Chronic lower respiratory disease (ORR 0.670-0.798), non-B & non-C hepatitis (ORR 0.703-0.765), lower digestive system diseases (ORR 0.698-0.820), dementia (ORR 0.573-0.656, and neurosis (ORR 0.642-0.807) showed a decreased risk of COVID-19 in multivariable analysis. This pattern was consistent in the subgroup which excluded long term health care utilizers (Table S9).

### Baseline demographics between severe and non-severe confirmed COVID-19

severe cases accounted for 5.6% (293 of 5,172) of the laboratory confirmed cases. The mean age was 65 years (range 18-94) and 52.0% (153 of 293 patients) were male in the severe group. The severe group had older patients and the male ratio was higher than that in the non-severe group. The proportion of Daegu/Gyeongbuk area (DG area, 60.4%, 177 of 293), median CCI, the number of healthcare utilization, and the prevalence of comorbidities were higher in severe group than that of non-severe group (Table 2).

**Table 2.** Baseline demographics and Prevalence of comorbidities between severe and non-severe laboratory confirmed COVID-19

| Variables | Severe<br>(N=293) | Non-Severe<br>(N=4,879) | P-value |
| --- | --- | --- | --- |
| **Demographic characteristics** |  |  |  |
| Sex, Male, n (%) | 153 (52.2) | 2,136 (43.8) | 0.0047 |
| Age, mean (range) | 65 (18-94) | 43 (18-100) |  |
| 18-50, n (%) | 40 (13.7) | 3,174 (65.1) | <.0001 |
| 50-65, n (%) | 97 (33.1) | 1,047 (21.5) |  |
| 65-80, n (%) | 99 (33.8) | 509 (10.4) |  |
| ≥80, n (%) | 57 (19.5) | 149 (3.1) |  |
| **Residence** |  |  |  |
| DG area, n (%) | 177 (60.4) | 1,158 (23.7) | <.0001 |
| Except DG, n (%) | 116 (39.6) | 3,721 (76.3) |  |
| Charlson comorbidity index, mean (SD) | 2.57 (2.4) | 1.27 (1.9) | <.0001 |
| **Healthcare utilization within 1 years before diagnosis of COVID-19** |  |  |  |
| Number of hospitalization, mean (SD) | 0.69 (1.3) | 0.48 (1.6) | 0.0090 |
| Number of hospitalization over 30 days, mean (SD) | 0.02 (0.2) | 0.01 (0.1) | 0.1788 |
| Number of outpatient visit, mean (SD) | 23.67 (34.1) | 24.83 (31.6) | 0.0175 |
| Number of ED visit, mean (SD) | 33.41 (40.1) | 23.08 (33.7) | <.0001 |
| Medical Aids, n (%) | 36 (12.3) | 251 (5.1) | <.0001 |
| **Underlying Diseases** |  |  |  |
| **Endocrinopathy** |  |  |  |
| Diabetes | 105 (35.8) | 608 (12.5) | <.0001 |
| Thyroid disease | 26 (8.9) | 299 (6.1) | 0.0600 |
| Cushing syndrome | 0 (0.0) | 10 (0.2) | 0.4380 |
| Osteoporosis | 54 (18.4) | 352 (7.2) | <.0001 |
| **Cardiac disease** |  |  |  |
| Isolated Hypertension | 156 (53.2) | 970 (19.9) | <.0001 |
| Ischemic heart disease | 44 (15.0) | 253 (5.2) | <.0001 |
| Heart failure and cardiomyopathy | 44 (15.0) | 173 (3.5) | <.0001 |
| Valvular heart disease | 1 (0.3) | 29 (0.6) | 0.5796 |
| Cardiac arrhythmia | 23 (7.8) | 171 (3.5) | 0.0001 |
| **Chronic respiratory disease** |  |  |  |
| Chronic upper respiratory disease | 187 (63.8) | 3,441 (70.5) | 0.0149 |
| Chronic lower respiratory disease | 131 (44.7) | 1,401 (28.7) | <.0001 |
| Environmental lung disease | 1 (0.3) | 9 (0.2) | 0.5529 |
| Interstitial lung disease | 4 (1.4) | 14 (0.3) | 0.0023 |
| Chronic respiratory failure and | 0 (0.0) | 4 (0.1) | 0.6240 |
| Diaphragm palsy |  |  |  |
| Pulmonary vascular disease | 1 (0.3) | 14 (0.3) | 0.8666 |
| **Renal disease and ESRD** |  |  |  |
| Hypertensive renal disease | 4 (1.4) | 57 (1.2) | 0.7617 |
| Glomerular disease | 13 (4.4) | 108 (2.2) | 0.0145 |
| Renal tubule-interstitial disease | 3 (1.0) | 28 (0.6) | 0.3325 |
| History of Acute Renal failure | 1 (0.3) | 26 (0.5) | 0.6585 |
| Chronic renal failure and ESRD | 17 (5.8) | 217 (4.4) | 0.2787 |
| Urolithiasis | 4 (1.4) | 82 (1.7) | 0.6817 |
| **Viral hepatitis and Chronic liver disease** |  |  |  |
| HBV, acute and chronic | 7 (2.4) | 93 (1.9) | 0.5599 |
| HCV, acute and chronic | 2 (0.7) | 11 (0.2) | 0.1291 |
| Non-B, non-C hepatitis | 50 (17.1) | 400 (8.2) | <.0001 |
| Liver cirrhosis | 2 (0.7) | 38 (0.8) | 0.8551 |
| Hepatic failure | 0 (0.0) | 7 (0.1) | 0.5165 |
| **Disease of digestive system** |  |  |  |
| Non-infectious disease of Upper Digestive system | 276 (94.2) | 4,528 (92.8) | 0.3680 |
| Non-infectious disease of Lower Digestive system | 153 (52.2) | 1,685 (34.5) | <.0001 |
| Pancreatic disease | 1 (0.3) | 56 (1.1) | 0.1991 |
| Biliary disease | 17 (5.8) | 107 (2.2) | <.0001 |
| **Chronic neurologic disease** |  |  |  |
| Systemic atrophy | 0 (0.0) | 5 (0.1) | 0.5836 |
| Parkinsonism and movement disorder | 25 (8.5) | 184 (3.8) | <.0001 |
| Alzheimer and degenerative disease | 23 (7.8) | 111 (2.3) | <.0001 |
| Multiple sclerosis | 0 (0.0) | 1 (0.0) | 0.8064 |
| Epilepsy | 10 (3.4) | 87 (1.8) | 0.0458 |
| Transient cerebral ischemia | 58 (19.8) | 291 (6.0) | <.0001 |
| Stroke, Cerebral hemorrhage |  |  |  |
| Dementia | 46 (15.7) | 126 (2.6) | <.0001 |
| **Malignancy** |  |  |  |
| Solid organ, except respiratory, thyroid | 25 (8.5) | 222 (4.6) | 0.0019 |
| Respiratory tract | 4 (1.4) | 29 (0.6) | 0.1076 |
| Thyroid cancer | 6 (2.0) | 61 (1.3) | 0.2410 |
| Hematologic | 1 (0.3) | 16 (0.3) | 0.9690 |
| **Rheumatologic disease** |  |  |  |
| Rheumatoid arthritis | 17 (5.8) | 100 (2.0) | <.0001 |
| SLE | 0 (0.0) | 0 (0.0) |  |
| Systemic connective tissue disease | 5 (1.7) | 46 (0.9) | 0.1989 |
| **Hematologic disease** |  |  |  |
| Anemia | 53 (18.1) | 537 (11.0) | 0.0002 |
| Coagulopathy | 3 (1.0) | 39 (0.8) | 0.6775 |
| Bone marrow dysfunction | 3 (1.0) | 42 (0.9) | 0.7704 |
| **Obesity** | 0 (0.0) | 2 (0.0) | 0.7289 |
| **Nutritional deficiency** | 21 (7.2) | 218 (4.5) | 0.0326 |
| **Mental and Behavioral disorders** |  |  |  |
| Mental disorder of Substance use | 6 (2.0) | 30 (0.6) | 0.0042 |
| Schizophrenia | 21 (7.2) | 97 (2.0) | <.0001 |
| Mood disorder | 60 (20.5) | 528 (10.8) | <.0001 |
| Neurosis | 75 (25.6) | 745 (15.3) | <.0001 |
| Personality disorder | 1 (0.3) | 10 (0.2) | 0.6227 |
| Mental retardation, Development disorder | 1 (0.3) | 16 (0.3) | 0.9690 |
| **Immune deficiency, HIV infection** | 0 (0.0) | 9 (0.2) | 0.4619 |

DG, Daegu city and Gyeongsangbuk-do province area. ESRD, end-stage renal disease. HBV, hepatitis B virus.
HCV, hepatitis C virus. SLE, systemic lupus erythematosus. HIV, human immunodeficiency virus.

### Comorbidities associated with severity of COVID-19

#### Result of Overall analysis

The ORs for the severity of COVID-19 according to the 56 categories of comorbidity are shown in Figure 4 and Table S10. Most disease category except thyroid disease, environmental lung disease, CRF and ESRD, hepatitis B viral disease, pancreatic disease, respiratory track malignancy were associated with an increased risk of severe COVID-19 in the univariate analysis. No-infectious lower digestive system disease (ORR 1.361-1.418) and schizophrenia (ORR 3.343-3.833) were significantly associated with an increased risk of severe COVID-19 in all multivariable analysis. Isolated hypertension (OR 1.378, 95% CI 1.008-1.883), rheumatoid arthritis (ORR 1.865-1.908), and substance use (ORR 2.790-2.848) showed significant association with the severity of COVID-19 in multivariable model 1, while heart failure and cardiomyopathy (ORR 1.562-1.730), chronic lower respiratory diseases, interstitial lung disease (ORR 1.361-1.413) showed significant association in multivariable model 2. CRF and ESRD (ORR 0.222-0.312) showed negative association with severity of COVID-19 in multivariable analysis (Figure 4, Table S10). In terms of the subgroup analysis which excluded long term health care utilizers, diabetes showed an increased risk of severe disease in multivariable model 2 (ORR 1.411-1.416) (Table S11).

**Figure 4.**
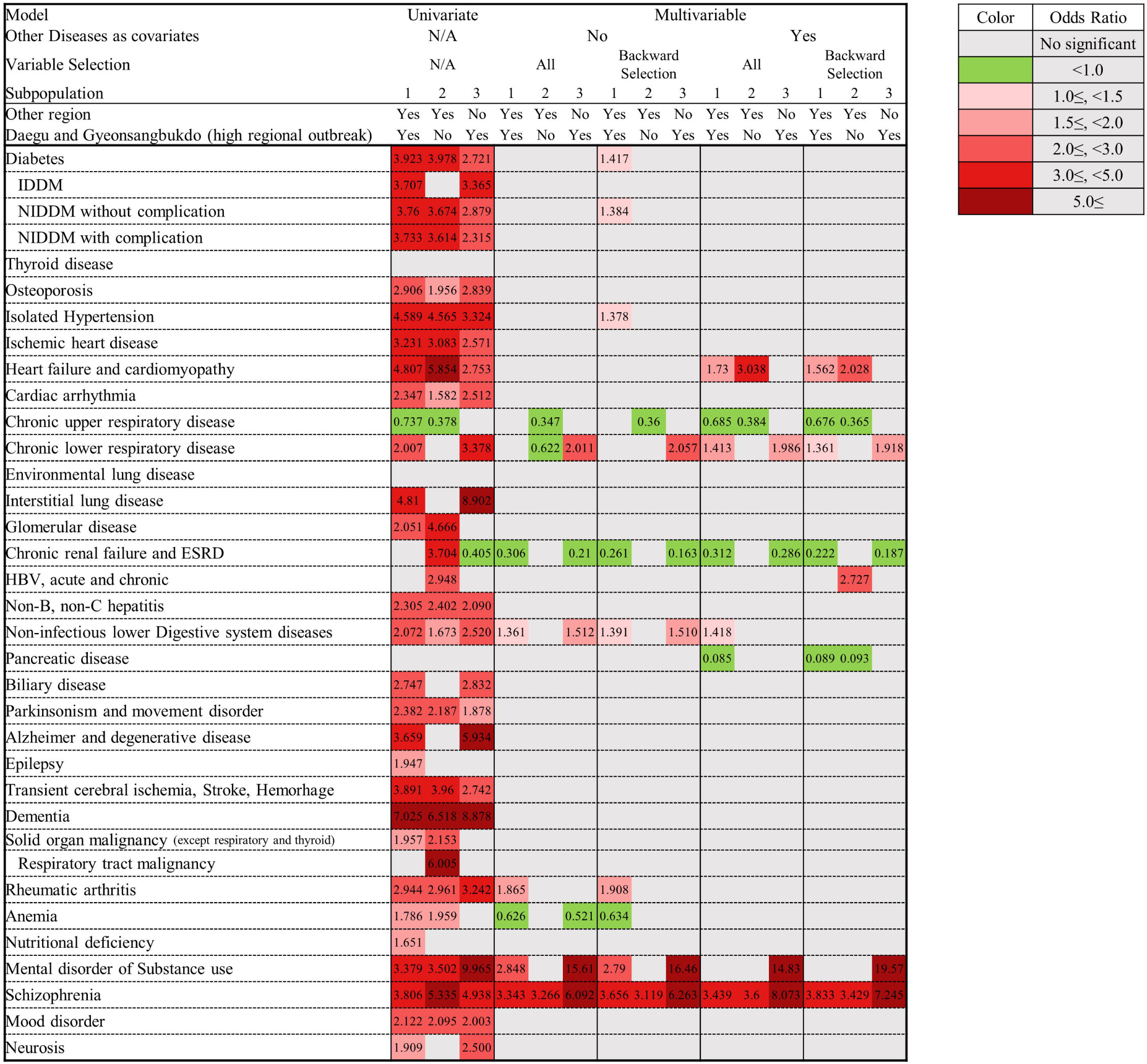
Analysis of relationship between comorbidities on severity of COVID-19.

#### Analysis of subpopulation which excluded high regional outbreak area (DG area)

In multivariable analysis, schizophrenia (ORR 3.119-3.600) was significantly associated with severe COVID-19, while chronic upper respiratory disease (ORR 0.347-0.384) showed a decreased risk of severe disease. Moreover, heart failure and cardiomyopathy (ORR 2.028-3.038), and hepatitis B viral disease (OR 2.727, 95% CI 1.032-7.202) showed an increased risk of severe disease only in multivariable model 2 (Table S12). This pattern was consistent in the subgroup which excluded long term health care utilizer (Table S13).

#### Analysis of subpopulation in high regional outbreak area (DG area)

In multivariable analysis, chronic respiratory disease (ORR 1.918-2.057), substance use (ORR 14.830-19.570), and schizophrenia (ORR 6.092-8.073) were significantly associated with risk of severe COVID-19. Non-infectious lower digestive system disease (ORR 1.510-1.512) showed an increased risk of severe disease in multivariable model 1 (Table S14). In terms of subgroup analysis except the long term health care utilizer, chronic upper respiratory disease showed an increased risk of severe COVID-19 in multivariable model 1 with backward selection (OR 1.515, 95% CI 1.010-2.271) (Table S15).

## DISCUSSION

In this study, we identified the possible comorbidities that might be associated with the risk of COVID-19 infection and its severity. Cushing syndrome, chronic renal disease, anemia, bone marrow dysfunction, and schizophrenia showed significant association with the diagnosis of COVID-19. The patterns of comorbidities associated with occurrence of COVID-19 might be different between high and non-high regional outbreak area. Chronic renal disease, solid organ malignancy, anemia, and bone marrow dysfunction were associated with community outbreak, whereas thyroid cancer, schizophrenia, and neurosis were associated with containment area. Diabetes, isolated hypertension, heart failure, chronic lower respiratory disease, non-infectious lower digestive system disease, rheumatoid arthritis, substance use, and schizophrenia were associated with severe COVID-19. Moreover, schizophrenia was strongly associated with diagnosis and severity of COVID-19.

When interpreting the results of this study, it is necessary to consider the demographic differences between the case and control group. Most diseases, except hypertensive renal disease, showed a higher prevalence in the control group compared to the case group (Table 1). This suggested that more tests for COVID-19 were done in people at risk of febrile respiratory illness regardless of COVID-19 pandemic[8-10]. Therefore, we presumed that the negative association in univariate analysis related to COVID-19 occurrence was based on biased baseline demographics and found that this selection bias was alleviated in the multivariate analysis (Figure 3). Moreover, we compared the laboratory confirmed case to the RT-PCR negative control groups, and conducted subpopulation analysis according to high regional outbreak areas where this bias would have been less affected. As a result, the comorbidities that increased the risk of diagnosis of COVID-19 were more meaningful. Thus, we found that Cushing syndrome, chronic renal diseases, anemia, bone marrow dysfunction, diabetes with complications, and solid organ malignancy might be risk factors for the diagnosis of COVID-19. In the case of thyroid cancer and mood disorder, consideration as risk factors was unjustifiable. However, schizophrenia was shown as a risk factor for COVID-19 in the overall and subpopulation analysis in the non-high regional outbreak area. Some early case reports about COVID-19 outbreak suggested that, differences in socioeconomic levels may affect SARS-CoV-2 infection[11]. It might suggest that the accompanying low socioeconomic status might increase the risk of developing COVID-19 rather than the schizophrenia itself[12].

In spite of a potential selection bias, the result suggested the possibility of hypertension, chronic lower respiratory disease, liver cirrhosis, lower digestive system diseases, pancreatic disease, biliary disease, epilepsy, dementia, obesity, and mood disorder being associated with a decreased risk of COVID-19. The patients with these conditions might have reduced social activity which may reduce the possible risk of exposure to SARS-CoV-2 virus. However, this is not clear, and more precise research is needed.

In early epidemiological studies of COVID-19, it was reported that comorbidities including diabetes, hypertension, and chronic respiratory disease were related to disease severity or death[4, 5, 13-15] as seen in this study except for malignancy. We found that heart failure and cardiomyopathy were associated with the severity of COVID-19. Human angiotensin-converting enzyme 2 (ACE2) is a functional receptor of SARS-CoV-2[16] and the heart is a tissue rich in ACE receptors along with the lungs. Cases of COVID-19 related to myocarditis were reported in China[17] and South Korea[18], and also reported as a risk factor for severe COVID-19[3].

This study also found that, non-infectious lower digestive system disease including inflammatory bowel disease and rheumatoid arthritis were only associated with the severity of COVID-19. Although the impact of immunosuppression on the severity of COVID-19 remains unclear, it is suggested that the long-term use of steroid or immunosuppressants may increase disease severity. Substance use and schizophrenia also showed a significant increasing risk of severe COVID-19; however, this may be caused by the low socio-economic status. Chronic upper respiratory disease appeared to be a risk factor for severe disease only in the community outbreak area, and decreased the risk in the other region. Lack of medical resources in the place where community outbreak occurred, led to denied proper medical management in patients with early respiratory symptoms, therefore increasing their risk for severe disease. In contrast, the patients who lived in the non-high regional outbreak area received proper management of their upper respiratory symptoms in the early phase of disease[19, 20]. For the same reason it might explain that the increased risk of presence of COVID-19 and the decreased risk of severe disease showed in patient with ESRD who received regular dialysis treatment. Because a patients with ESRD are known to be vulnerable to infectious disease, while they might be likely to receive early appropriate medical treatment suggested in previous literature[21].

This study had a few limitations. Firstly, this study was limited to data from the nationwide claim database of subjects who received laboratory test of COVID-19. Thus, the database including those who were tested via “Drive Through” or local health centers and treated in non-medical facility was not included in this study. Therefore, the number of officially announced population was different from that of the analyzed population, which was used in this study. Despite this limitation, this study was conducted only for those who performed the laboratory test for COVID-19 on insurance claims database. Therefore, the negative control group had a confirmed a negative result of SARS-CoV-2 infection. Thus, the comparison between this negative control group and case group helped in proper evaluation of the risk factors for COVID 19 occurrence. Another limitation was the inability of the data source to provide the information on the severity of the comorbidities. Finally, we could not evaluate the detailed mechanism of the relationship between comorbidities and the diagnosis or severity of COVID-19. However, most comorbidities identified in each individual’s health insurance claim data could be used for this study. It could be possible to discover previously unknown or unexpected risk factors based on a data driven approach.

## CONCLUSION

In this retrospective case control study, we suggested that Cushing syndrome, chronic renal diseases, anemia, bone marrow dysfunction, diabetes with complications, and solid organ malignancy might be the risk factors for COVID-19. Besides, diabetes, hypertension, chronic respiratory disease, heart failure and cardiomyopathy, non-infectious lower digestive system disease, rheumatoid arthritis, and mental disorder owing to substance use were related with severe COVID-19 disease. In addition, schizophrenia was associated with the occurrence and severity of COVID-19.

## Data Availability

You can check the data on the site below.
https://hira-covid19.net/

## STATEMENTS

## Acknowledgment

The authors appreciate healthcare professionals dedicated to treating COVID-19 patients in Korea, and the Ministry of Health and Welfare and the Health Insurance Review & Assessment Service of Korea for sharing invaluable national health insurance claims data in a prompt manner.

## Statement of Ethics

The research was conducted ethically in accordance with the World Medical Association Declaration of Helsinki and was approved by the appropriate institutional review board of the *Gachon University College of Medicine, Incheon*, Republic of Korea (GFIRB2020-118).

## Author Contribution

Study concept and design: WJ, KH, JJ, MK

Data collection and conducting procedures: MK. JH, GHB, RL, YN, HC, Y-HC, SG

Data analysis and interpretation: WJ, KH, JJ, K-PK, J-SI

Drafting of the manuscript: WJ, KH, JJ

Critical revision of the manuscript: WJ, KH, JJ

## Online Supplementary Materials

**Table S1. Variables in the Korean National Health Insurance claim data**.

**Table S2. ICD 10 codes for underlying comorbidities category**

**Table S3. International Classification of Diseases 10th Revision (ICD-10) mapping for Charlson Comorbidity Index**

**Table S4. Analysis of relationship between underlying disease and diagnosis of COVID-19 in whole study participants (Anlysis1)**

**Table S5. Subgroup analysis of relationship between underlying disease and diagnosis of COVID-19 except the participants who had the history of hospitalization over 30 days prior to diagnosis (Anlysis5)**

**Table S6. Subgroup analysis of relationship between underlying disease and diagnosis of COVID-19 except the Daegu/Gyeongbuk region (Anlysis3)**

**Table S7. Subgroup analysis of relationship between underlying disease and diagnosis of COVID-19 except the Daegu/Gyeongbuk region and the participants who had the history of hospitalization over 30 days prior to diagnosis (Anlysis7)**

**Table S8. Subgroup analysis of relationship between underlying disease and diagnosis of COVID-19 in the Daegu/Gyeongbuk region (Anlysis9)**

**Table S9. Subgroup analysis of relationship between underlying disease and diagnosis of COVID-19 in the Daegu/Gyeongbuk region, but except the participants who had the history of hospitalization over 30 days prior to diagnosis (Anlysis11)**

**Table S10. Analysis of relationship between underlying disease and severity of COVID-19 in whole study participants (Anlysis2)**

**Table S11. Subgroup analysis of relationship between underlying disease and severity of COVID-19 except the participants who had the history of hospitalization over 30 days prior to diagnosis (Anlysis6)**

**Table S12. Subgroup analysis of relationship between underlying disease and severity of COVID-19 except the Daegu/Gyeongbuk region (Anlysis4)**

**Table S13. Subgroup analysis of relationship between underlying disease and severity of COVID-19 except the Daegu/Gyeongbuk region and the participants who had the history of hospitalization over 30 days prior to diagnosis (Anlysis8)**

**Table S14. Subgroup analysis of relationship between underlying disease and severity of COVID-19 in the Daegu/Gyeongbuk region (Anlysis10)**

**Table S15. Subgroup analysis of relationship between underlying disease and diagnosis of COVID-19 in the Daegu/Gyeongbuk region, but except the participants who had the history of hospitalization over 30 days prior to diagnosis (Anlysis12)**

